# Procalcitonin and High APACHE Scores are Associated with the Development of Acute Kidney Injury in Patients with SARS-CoV-2

**DOI:** 10.1101/2022.08.09.22274874

**Authors:** Andrade Sierra Jorge, Delgado Astorga Claudia, Nava Vargas Miriam Gabriela, Rojas Campos Enrique, Arellano Arteaga Kevin Javier, Hernández Morales Karla, Carlos A Andrade Castellanos, Andrade-Ortega Antonio de Jesús, González-Correa Luis Gerardo

**Author notes:** **Corresponding author:** Jorge Andrade Sierra, MD, PhD, Department of Physiology, University Health Sciences Center (*Centro Universitario de Ciencias de la Salud*), University of Guadalajara, Guadalajara, Jalisco, Mexico. Sierra Mojada No. 950, Col. Independencia, Guadalajara, Jalisco, México CP 44340.

## Abstract

**Background:** Acute kidney injury (AKI) is associated with poor outcomes in patients infected with SARS-CoV-2. Sepsis, direct injury to kidney cells by the virus, and severe systemic inflammation are mechanisms implicated in its development. We investigated the association between inflammatory markers (C-reactive protein, procalcitonin, D-dimer, lactate dehydrogenase, and ferritin) in patients infected with SARS-CoV-2 and the development of AKI.

**Methods:** A prospective cohort study performed at the Civil Hospital (Dr. Juan I. Menchaca) Guadalajara, Mexico, included patients aged >18 years with a diagnosis of SARS-CoV-2 pneumonia confirmed by RT-PCR and who did or did not present with AKI (KDIGO) while hospitalized. Biomarkers of inflammation were recorded, and kidney function was estimated using the CKD-EPI formula.

**Results:** 291 patients were included (68% men; mean age, 57 years). The incidence of AKI was 40.5% (118 patients); 21% developed stage 1 AKI, 6% developed stage 2 AKI, and 14% developed stage 3 AKI. The development of AKI was associated with phosphate higher (p = 0.002) (RR 1.39, CI 95% 1.13 – 1.72), high procalcitonin levels at hospital admission (p = 0.005) (RR 2.09, CI 95% 1.26-3.50), and high APACHE scores (p = 0.011) (RR 2.0, CI 95% 1.17-3.40). The survival analysis free of AKI according to procalcitonin levels and APACHE scores demonstrated a lower survival in patients with procalcitonin >0.5 ng/ml (p= 0.001) and APACHE >15 points (p = 0.004).

**Conclusions:** phosphate, high procalcitonin levels, and APACHE scores >15 were predictors of AKI development in patients hospitalized with COVID-19.

## Introduction

The coronavirus 19 (COVID-19) pandemic, which originated with the novel coronavirus (SARS-CoV-2), caused 318 thousand deaths in Mexico as of February 2022 (1). Older adults with comorbidities(2, 3) are at a higher risk of complications from SARS-CoV-2 infections. Kidney damage is one of the main complications and can be demonstrated by the presence of hematuria, proteinuria (4-6), and the development of acute kidney injury (AKI), with high incidence reported in hospitalized patients.(7-10) The severity leads to even higher mortality, which has a multifactorial etiology (11, 12). Nevertheless, baseline characteristics, patient interventions in critical care, and organ crosstalk are mechanisms that influence the appearance of AKI, but possible direct injury by the virus on kidney cells (podocytes, proximal tubule cells, and the epithelial cells of Bowman’s capsule (13-15)) and uncontrolled systemic inflammation are factors that also influence its development and severity in patients with COVID-19 (4, 16, 17). Inflammation and the consequent increase in some biomarkers also appear to be associated with kidney damage and poor outcomes (2, 3, 18-25). In fact, the therapeutic conduct for the prevention of developing AKI associated with COVID-19 is similar to other etiologies (avoiding nephrotoxins, periodic review of serum creatinine (SCr), urinary output, and hemodynamic monitoring), and in critically ill patients with COVID-19, it could reduce the appearance or severity of AKI (26). Our hospital has been a referral center for patients without social security throughout the pandemic, and the limitations of some resources, such as urinary markers (neutrophil gelatinase-associated lipocalin (NGAL) and kidney injury molecule 1 (KIM-1)), demand the use of low-cost inflammatory markers to predict the development of AKI. Our objective was to determine the existing association between CRP, PCT, D-dimer, LDH, and ferritin levels and the development and severity of AKI in patients hospitalized with pneumonia caused by SARS-CoV-2 without social security.

## Methods

A prospective cohort study was performed in patients without social security coverage (Internal Medicine Department “Dr. Juan I. Menchaca” Civil Hospital in Guadalajara) from March 1, 2020, to March 1, 2021. All patients tested positive for SARS-CoV-2 using reverse transcriptase polymerase chain reaction (RT-PCR). Based on the development of AKI using the Kidney Disease Improving Global Outcomes (KDIGO) classification, patients were classified into two groups: those with and without AKI. All patients in our study had normal kidney function when admitted to the hospital, and we could not clinically determine infections other than COVID-19.

### Data collection

Quantitative polymerase chain reaction (RT-PCR) was used to determine the presence of SARS-CoV-2 in the nasopharyngeal swabs. All registered patients were tested for inflammatory biomarkers (lactate dehydrogenase, ferritin, highly sensitive C-reactive protein, D-dimer, and procalcitonin) upon admission and during hospitalization, and the following data were collected: age, sex, history of smoking, alcoholism, prescribed medication use, and comorbidities. The patients’ anthropometric characteristics and presence of other infections (as documented by clinical evaluation or confirmed by any microbiological method) were also recorded. The following biochemical variables were recorded: glucose, urea, SCr, uric acid, glycated hemoglobin, lipids, liver enzymes, electrolytes, hemoglobin, platelets, leukocytes, and lymphocytes. Imaging studies, such as chest radiography or tomography, were performed on all patients upon hospital admission. Disease severity was assessed using the Acute Physiology and Chronic Health Evaluation II (APACHE II) scoring system and sequential organ failure assessment (SOFA). Glomerular filtration rate (eGFR) was estimated using the CKD-EPI formula (27).

Patients with chronic kidney disease (CKD), kidney transplant, obstructive kidney comorbidities, single kidney, neoplastic or autoimmune diseases, chronic use of anti-inflammatory drugs, and use of immunosuppressant drugs were excluded from the study The present study complies with the ethical principles for medical research in human beings as stipulated by the Declaration of Helsinki, 64th General Assembly, Fortaleza, Brazil, October 2013, in addition to adhering to the standards of good clinical practice. All procedures were performed according to the national regulations stipulated in the General Health Legal Guidelines for Health Care Research in Mexico, 2nd Title, Ethical Aspects for Research in Human Beings, Chapter 1, Article 17. The study protocol was reviewed and approved by the Ethics Committee of the Hospital Civil de Guadalajara, “Dr. Juan I Menchaca” (*HCG-JIM*). Registration number: 17CI14 039 116 COFEPRIS. Guadalajara, Jalisco, México.

## Consent to participate statement

All patients signed informed consent forms prior to hospital admission and provided consent to participate in the study.

### Inflammatory markers

#### Procalcitonin

Measurements were performed using the sandwich principle of an automated electrochemiluminescent immunoassay (Cobas E411,Roche).

#### Ferritin

Measured using the immunoturbidimetric assay: Image 800 (Dyasys).

#### Highly sensitive C-reactive protein (hsCRP)

Measurements were performed using a particle-enhanced immunoturbidimetric assay Cobas® c501 (Roche).

#### D-Dimer

Measured using an immunochromatography assay: Ramp®.

### Definitions

#### SARS CoV-2 pneumonia

defined by the National Institutes of Health as individuals with SARS-CoV-2 infection confirmed by RT-PCR testing who have SpO_2_ <94% on room air at sea level, a ratio of arterial partial pressure of oxygen to fraction of inspired oxygen (PaO_2_/FiO_2_) <300 mmHg, respiratory frequency >30 breaths/min, or lung infiltrates >50% (28).

#### Acute kidney injury (AKI) (29)

An abrupt decrease in GFR manifested by an increase in SCr or oliguria within the first 48 hours to seven days.

We classified AKI considering only the increase in SCr:

AKI1: Increase in SCr 1.5 to 1.9 times the baseline level, or an increase of >0.3 mg/dl.

AKI2: Increase in SCr 2 to 2.9 times the baseline level.

AKI3: Increase in SCr by 3 times the baseline, an increase of SCr >4 mg/dl, or the onset of renal replacement therapy.

### Statistical analysis

The data are presented as mean ± standard deviation or median, numbers, and percentages where appropriate. Student’s t-test or the Mann-Whitney U test, depending on the distribution, was used to compare groups. The Cox proportional hazards model was used to estimate the risk of AKI. Survival free of AKI and mortality was evaluated using the Kaplan-Meier test. Statistical analysis was performed using SPSS™ software, version 17. Statistical significance was set at *p*<0.05.

## Results

A total of 291 patients were eligible for inclusion in this study. The average age was 57±14 years, and 68% (n=198) corresponded to the male sex. At the time of hospital admission, the average body mass index (BMI) was 31 kg/m2, and approximately 40% of the patients had an associated comorbidity (DM or SAH). Ninety-five percent (95%) of the patients required some form of supplementary oxygen, and 20% were categorized as having severe illness requiring high-flow nasal cannulas and/or invasive mechanical ventilation (IMV) with an average of 9.5 days of hospitalization. On admission, no infectious processes other than the infection due to COVID-19, and all patients received dexamethasone at a dosage of 6 mg/day for 10 days during their hospital stay. The baseline characteristics of the patients are presented in TABLE 1.

**Table 1.** Demographic characteristics of patients diagnosed with acute kidney injury (AKI) and COVID-19.

| Characteristics | Total<br>n=291 | AKI<br>n=118 | No AKI<br>n=173 |
| --- | --- | --- | --- |
| Age (years) | $57 \pm 14$ | $61 \pm 14^*$ | $54 \pm 13^*$ |
| Sex- Masculine, n (%) | 198 (68) | 88 (75) * | 110 (64) * |
| Smoking, n (%) | 89 (31) | 41 (35) | 48 (28) |
| Hospitalization (days) | $9.5 \pm 7.0$ | $12 \pm 7.8^*$ | $7.7 \pm 5.7^*$ |
| Comorbidities |  |  |  |
| BMI, ($\text{Kg/m}^2$) | $30.9 \pm 6.9$ | $31.2 \pm 7.5$ | $30.6 \pm 6.4$ |
| Diabetes, n (%) | 113 (39) | 55 (46.6) * | 58 (33.5) * |
| Hypertension, n (%) | 112 (38) | 58 (49.2) * | 54 (31.2) * |
| Oxygen requirement n (%) |  |  |  |
| No use | 5 (1.7%) | ---- | 5 (2.9%) |
| Nasal prongs | 94 (32.3%) | 30 (25.5%) | 64 (37%) |
| Mask | 133 (45.7%) | 49 (41.5%) | 84 (48.6%) |
| High flow | 42 (14.4%) | 28 (23.7%) | 14 (8 %) |
| CPAP | 1 (0.3%) | ---- | 1 (0.6) |
| IMV | 16 (5.5%) | 11 (9.3%) | 5 (2.9) |
| **Medications use** |  |  |  |
| Metformin, n (%) | 72 (25) | 38 (32.2) * | 34 (19.7) * |
| ACEi or ARBs, n (%) | 75 (26) | 25 (21) | 50 (29) |
| Diuretics, n (%) | 51 (18) | 44 (37) * | 7 (4) * |
| Dexamethasone, n (%) | 285 (97.9) | 116 (98.3) | 169 (98) |
| **Evaluation of scales** |  |  |  |
| APACHE II (pts) | 9.70 ± 5.43 | 12.7 ± 6.0* | 7.7 ± 3.9* |
| SOFA (pts) | 3.06 ± 2.40 | 4.1 ± 3.0* | 2.40 ± 1.50* |
| **Laboratory exams at admission** |  |  |  |
| Hemoglobin (gr/dL) | 14.65 ± 2.30 | 14.4 ± 2.4 | 14.8 ± 2.2 |
| Leukocytes (x10<sup>9</sup>/L) | 11.16 ± 5.21 | 12.6 ± 5.58 | 10.0 ± 4.7 |
| Lymphocytes (x10<sup>9</sup>/L) | 9.73 ± 5.17 | 0.94 ± 0.52 | 1.0 ± 0.51 |
| Platelets (x10<sup>9</sup>/L) | 263 ± 89 | 266 ± 91 | 260 ± 87 |
| Glucose (mg/dL) | 183 ± 121 | 215 ± 151* | 161 ± 88* |
| Hemoglobin A1c (%) | 7.9 ± 2.5 | 8.18 ± 2.6 | 7.8 ± 2.4 |
| **Inflammatory markers at admission** |  |  |  |
| LDH (u/L) | 391 ± 169 | 428 ± 166 | 367 ± 168 |
| Ferritin (µg/L) | 815 ± 529 | 884 ± 592 | 737 ± 486 |
| D-Dimer (mg/mL) | 434(170-1483) | 670(230- 2706)* | 327(148-927)* |
| CRP (mg/dL) | 148 ± 108 | 158 ± 110 | 142 ± 107 |
| Procalcitonin (ng/mL) | 0.28(0.11-.77) | 0.72(0.34-2.01)* | 0.16(0.09-.34)* |
| **Severe Sepsis/Septic Shock, n (%)** | 66 (23) | 58 (49) * | 8 (4.6) * |
| **Creatinine, at baseline (mg/dL)** | .80 ± 0.19 | 0.87 ± 0.22 | 0.74 ± 0.15 |
| **Stages of AKI, n (%)** |  |  |  |
| 1 | --- | 61(51.7%) | --- |
| 2 | --- | 16 (13.6 %) | --- |
| 3 | --- | 41 (34.7%) | --- |

| Recovery from AKI, n (%) |  |  |  |
| --- | --- | --- | --- |
| Yes | --- | 65 (55) | --- |
| No | --- | 53 (45) | --- |

**AKI:** acute kidney injury; **BMI:** body mass index; **CRP:** C-reactive protein; **LDH:** lactate dehydrogenase; **CPAP:** continuous positive airway pressure; **IMV:** invasive mechanical ventilation. **ACEi or ARBs:** angiotensin-converting enzyme inhibitors or angiotensin receptor blockers \* $p < 0.05$ Comparison between groups with and without AKI.

### Acute kidney injury (AKI)

AKI was recorded in 40.5% (118 of the 291 patients). Seventy-one patients (21%) had stage 1 AKI, 16 (6%) had stage 2 AKI, and 41 (14%) had stage 3 AKI. The development of severe sepsis or septic shock during follow-up was documented in 49% (58 of 118) *(p<0.05)*. In the group that developed AKI, male sex predominated (75% *vs*. 64% in the group without AKI, respectively), older age (61 *vs*. 54 years), and prolonged hospital stay (12 *vs*. 7.7 days) *(p<0.05)*. Approximately 50% of patients with AKI had some type of associated comorbidity (DM or SAH) compared to only 30% in the group without AKI *(p<0.05)*. The use of dexamethasone, angiotensin-converting enzyme inhibitors (ACEi), and angiotensin receptor blockers (ARBs) was similar in both groups, whereas the use of diuretics was higher in patients with AKI. The highest points in severity on the APACHE II and SOFA scores at the time of hospital admission were recorded in patients who developed AKI *(p<0.05)*. Baseline hemoglobin, platelets, leukocytes, lymphocytes, CRP, LDH, and ferritin levels at hospital admission did not differ between groups, while glucose levels (215 *vs*. 161), PCT (0.72 *vs*. 0.16), and D-dimer (670 mg/dL *vs*. 327 mg/dL) (p=0.001) were significantly higher in patients with AKI than in those without AKI. During follow-up, inflammatory markers in AKI stages 2 and 3 presented higher levels of PCT on days 6 and 12 (*p = 0.009 and p = 0.012*, respectively), ferritin on days 6 and 12 (p=NS), and D-dimer on day 6 (*p= 0.017*). TABLE 2

**Table 2.** Markers of inflammation on hospitalization and during follow-up in patients with COVID-19 and acute kidney injury (AKI).

|  | **AKI total**<br>**N = 118** | **AKI 1**<br>**N = 61** | **AKI 2 and 3**<br>**N = 57** |
| --- | --- | --- | --- |
| **Ferritin** |  |  |  |
| On hospitalization | 884±592 | 839±479 | 933±699 |
| Day-3 | 815±508 | 637±392 | 964±520 |
| Day-6 | 846±499 | 594±423 | 1111±439 |
| Day-12 | 972±510 | 781±516 | 1104±482 |
| **LDH (u/L)** |  |  |  |
| On hospitalization | 428±166 | 396±168 | 462±158 |
| Day-3 | 474±391<sup>§</sup> | 388±249 | 578±498 |
| Day-6 | 383±157 | 368±132 | 416±176 |
| Day-12 | 339±140 | 340±153 | 339±133 |
| **D-Dimer (ng/mL)** |  |  |  |
| On hospitalization | 670 (230-2706) | 543 (210-1830) | 970 (274-3260) |
| Day-3 | 1259 (472-4308) | 921(239-3683) | 2264 (855-2264) |
| Day-6 | 1727 (646-3911) | 960 (588-2534)* | 2928 (868-5000)* |
| Day-12 | 1782 (708-3201) | 1007 (461-1940) | 2234 (1702-4452) |
| **CRP (mg/dL)** |  |  |  |
| On hospitalization | 158±110 | 148±109 | 171±112 |
| Day-3 | 105±100<sup>§</sup> | 83±86 | 125±107 |
| Day-6 | 85±91 | 78±110 | 91±72 |
| Day-12 | 115±76 | 81±76 | 143±66 |
| **Procalcitonin (ng/mL)** |  |  |  |
| On hospitalization | 0.72 (0.34-2.01) | 0.53 (0.22-1.13)* | 1.0 (0.52-2.80)* |
| Day-3 | 0.42(0.20-1.90) | 0.28 (0.15-0.64)* | 1.13 (0.33-7.52)* |
| Day-6 | 0.49 (0.16-1.56) | 0.23 (0.11-.62)* | 1.29 (0.20-3.9)* |
| Day-12 | 1.18 (0.37-2.96) | 0.43 (0.15-1.16)* | 1.94 (0.56-6.10)* |

**AKI:** acute kidney injury; **CRP:** C-reactive protein; **LDH:** lactate dehydrogenase; § $p < 0.05$ vs. No AKI
\* $p = < 0.05$ AKI 1 vs. AKI 2 and 3.

In the multivariate analysis, phosphate higher (p = 0.002, RR 1.39, CI 95% 1.13 – 1.72), PCT level >0.5 ng/ml on hospitalization (*p = 0.005, RR 2.09, CI 95% 1.26-3.50*), and points >15 (*p = 0.011; RR 2.0 CI 95% 1.17-3.40*) showed a significant association with the development of AKI. TABLE 3 Fifty-five percent of those who developed AKI recovered kidney function, the majority of whom had AKI stage 1. The survival analysis free of AKI according to PCT levels (≥ 0.5 ng/ml) (*p = 0.001*) and points on the APACHE score (>15 points) (*p = 0.004*) demonstrated a lower survival. FIGURE 1.

**Table 3.** Multivariate analysis for risk factors associated with acute kidney injury (AKI) in patients with COVID-19.

| Variable | Multivariate Analysis |  |  |
| --- | --- | --- | --- |
|  | RR | CI 95% | P |
| Phosphate | 1.39 | 1.13 – 1.72 | 0.002 |
| APACHE | 2.0 | 1.17 – 3.40 | 0.011 |
| Procalcitonin | 2.09 | 1.26 – 3.50 | 0.005 |

**Figure 1.**
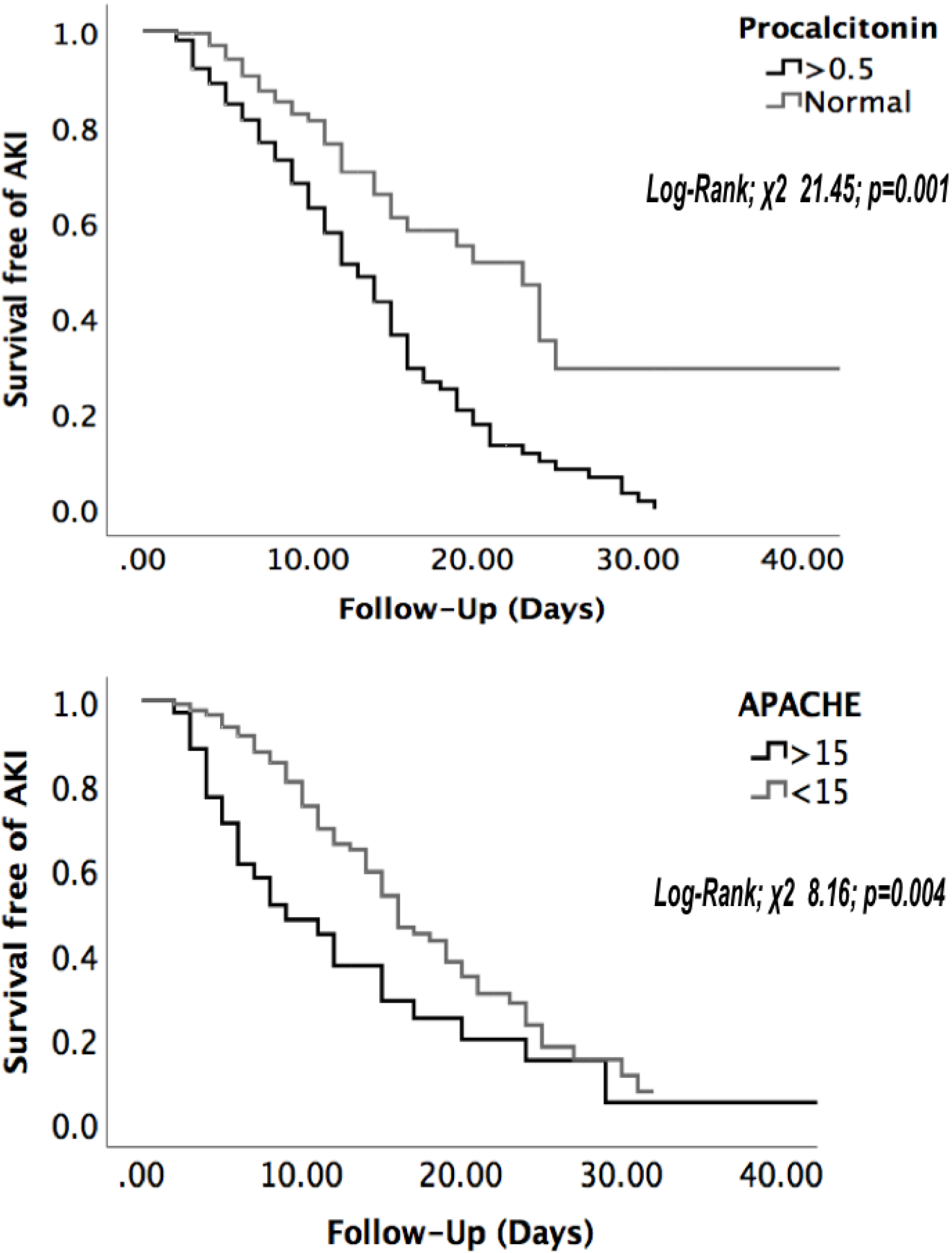
Acute kidney injury-free time according to procalcitonin and APACHE score.

## DISCUSSION

The present study concurs with the Latin American registry (30), which describes a high frequency of AKI (65%). In Mexico, incidences of 33.7 and 58.6% are reported, where half correspond to severe cases, with comorbidities consistent with the present study, where we documented that 30% of patients were overweight or had some degree of obesity.(31, 32) The development of AKI has been associated not only with obesity but also with being overweight (33). The present study did not perform an analysis of the obesity subgroups, but both study groups had similar BMIs, and there was no demonstrated association with the development of AKI. We found a high prevalence of SAH and DM with higher concentrations of glucose and HbA1C in those who developed AKI, but these were not associated with risk. Our results are consistent with the literature, where age is a risk factor for severe illness (19) and leads to a higher risk of AKI (34). Hirsch et al. demonstrated a higher risk of AKI (*OR 1.03, CI 95% 1.03-1.04; p < 0.001*) in those aged >60 years (7). Similarly, Fisher et al. (35) reported that older age (67 *vs*. 60 years; *p < 0.001*) was an independent risk factor for developing AKI. In Mexico, Casas-Aparicio *et al*. associated higher age with AKI development (*OR 1.07, CI 95% 1.01 - 1.13; p = 0.024*) (31). In our cohort, although the AKI group were older (61 ± 14 vs 54 ± 13; *p <0.001*), we were unable to consolidate these results not achieving statistical significance. Hyperphosphatemia is related to the risks of AKI and mortality(36). In this cohort we did not find hyperphosphatemia but but phosphathe levels were higher in AKI group. There is evidence that AKI is a phenomenon associated with the activation of proinflammatory cytokines, and with this, it calls for the use of markers of kidney injury (27). Some biomarkers predict AKI(37); however, the high costs and lack of availability of these tests in our region forced us to use biochemical parameters that are within reach in our practice to predict AKI events. PCT is barely detectable in healthy individuals; however, diverse conditions tend to elevate it (surgery, trauma, burns, pancreatitis, and sepsis), where levels can be markedly high (38-42). One of the advantages of PCT is its rapid increase 6 h after the onset of the inflammatory process (27). The elimination pathway of PCT is not clear, but due to its low molecular weight and deterioration of kidney function, it can elevate its levels; therefore, it is not a reliable parameter in the case of illness because the thresholds are different (43-45). Considering this aspect, the present study excluded patients with the onset of deterioration of kidney function and any patient who had conditions that would affect PCT values. The present multivariate analysis showed a significant risk for the development of AKI, with PCT rates >0.5 ng/ml at the moment of hospitalization. Some studies have demonstrated an association between PCT and the development of kidney damage in contexts other than COVID-19 (20, 38-40). Feng *et al*. showed the sensitivity (76%) and specificity (75%) of PCT in predicting AKI (*OR 9.63, 95% CI 95% 4.38–21.18*) (38), and Chun K. *et al*. (39) reported that in septic patients hospitalized in an intensive care unit (ICU), PCT was significantly high (*p <0.001*) and was associated with AKI (*OR 1.006, 1,000–1,011; p= 0,035*). In the present study, 50% of the patients who developed AKI had a case of severe sepsis or septic shock that developed during their hospital stay (p <0.05); however, our analysis did not demonstrate an association of the risk for the development of AKI. Regarding COVID-19, Wang *et al*. found that PCT levels were a predictor of kidney damage (46). This association can be measured by an exaggerated inflammatory response, even if the pathophysiological mechanism is unclear (47, 48). As a crucial component of the inflammatory cascade, the expression of PCT can be correlated with the levels of systemic inflammation and the development of AKI in patients with COVID-19 (63). Procalcitonin is a chemoattractant produced by monocytes in the inflammatory area, which contributes to the increase and recruitment of parenchymal cells with greater release of cytokines associated with acute kidney damage (39). Another implicated mechanism is the direct toxic effect of PCT on mesangial cells and apoptosis (48). Additionally, other markers demonstrate increments dependent on the reduction of the GFT (49), and in AKI, CRP and D-dimer levels have been shown to be high (31). The reduction in kidney function can explain our results of the increase in D-dimer levels predominantly in stages 2 and 3 of AKI; however, the analysis did not show an association with the risk for AKI. In severely ill patients, the APACHE II score is used over the long term to predict hospital mortality up to 28 days in different contexts (sepsis and pancreatitis). Zou et al. documented excellent discriminative power for mortality in patients with COVID-19 with point scores ≥17 (50). In the context of COVID-19, the APACHE II score was also associated with AKI in a Latin American cohort (*OR 1.97, CI 95% 1.08-2.64; p<0.05*), with higher mortality in those who had higher point scores (*OR 1.08, CI 95%, 1.02-1.98; p<0.05*) and in those who developed stage 3 AKI (*OR 1.11, CI 95%, 1.05-2.57; p<0.05*). These data support that in COVID-19 patients, AKI is a decisive phenomenon in the prognosis of these patients (67). In different studies on patients with COVID-19, higher APACHE II scores have been associated with poor prognosis and AKI severity (68, 69). We found that >15 points on the APACHE II score demonstrated significance for the development of AKI, which could be explained by the severity of the clinical scenario of COVID-19 and the patient’s comorbidities upon hospitalization. Until now, it remains unknown whether recovery from AKI in COVID-19 differs from other forms of AKI, and the direct, long-term impact of the SARS CoV-2 virus on kidney function in those who regain function remains unclear; however, the recommendation is a follow-up period of at least 2 to 3 months(51). Despite the high incidence of AKI, the majority of our patients had a favorable prognosis regarding recovery of kidney function, since more than half of the patients studied had an improvement in GFR; however, progression from stages 2 to 3 was associated with poor recovery.

## CONCLUSIONS

In this retrospective cohort, significantly high PCT levels were found on hospital admission in patients who developed AKI, and this was a predictor of AKI development. The trajectory of PCT throughout the follow-up did not show significant differences between those who did and did not develop AKI. Therefore, the use of PCT reinforces the importance of this marker as a predictor for the development of AKI in patients with COVID-19 infection without superinfections when admitted to the hospital.

## LIMITATIONS

The present study had some limitations. We recognize that this study is unicentric. However, being a referral center for patients infected with SARS-CoV-2 who do not have social security in our region favors extrapolation of the results to other populations, especially in those with limited access to health care. The majority of patients before hospitalization had passed a considerable amount of time since acquiring the viral infection; therefore, subclinical bacterial superinfection could not be ruled out. Economic limitations (lack of treatment with continuous slow therapies) could influence the lack of recovery of kidney function, especially in those who developed severe AKI. Finally, the lack of measurement and/or comparison of markers valid for AKI does not permit the determination of PCT levels.

## Data Availability

Most clinical data are restricted to protecting information from misuse. The data may be shared by the OPD, Hospital Civil de Guadalajara, and Dr. Juan I Menchaca (HCG-JIM) after approval from the Ethics Committee of the Institution. The data supporting the findings of this study are available from the corresponding author upon reasonable request.

## Abbreviations

COVID-19: Coronavirus 2019
SAH: Systemic arterial hypertension
DM: Diabetes mellitus
AKI: Acute kidney injury
CRP: C-reactive protein
PCT: Procalcitonin
LDH: Lactate dehydrogenase
ALT: Alanine aminotransferase
SCr: Serum creatinine
CRRT: Continuous Renal Replacement Therapy
RT-PCR: Reverse Transcriptase Polymerase Chain Reaction
KDIGO: Kidney Disease Improving Global Outcomes
APACHE II: Acute Physiology and Chronic Health Evaluation II
SOFA: Sequential Organ Failure Assessment
eGFR: Estimated glomerular filtration rate
CKD: Chronic Kidney Disease
PaO_2_/FiO_2_: Oxygen to fraction of inspired oxygen
BMI: Body Mass Index
IMV: Invasive mechanical ventilation
ACEi: Angiotensin-converting Enzyme Inhibitors
ARBs: Angiotensin receptor blockers
NGAL: Neutrophil gelatinase-associated lipocalin
KIM-1: Kidney injury molecule 1

## Data Availability

Most clinical data are restricted to protecting information from misuse. The data may be shared by the OPD, Hospital Civil de Guadalajara, and Dr. Juan I Menchaca (*HCG-JIM*) after approval from the Ethics Committee of the Institution. The data supporting the findings of this study are available from the corresponding author upon reasonable request.

## Conflict of interest

All the authors have declared no conflicts of interest.

## Funding statement

The study did not receive any private or governmental funding.

## Acknowledgements

Not applicable.

## Author Contributions

a. Conception and design for: Jorge Andrade-Sierra, Claudia Delgado Astorga and Enrique Rojas Campos.
b. Analysis and interpretation of data: Jorge Andrade-Sierra, Claudia Delgado Astorga, Miriam Gabriela Nava Vargas and Enrique Rojas Campos.
c. Drafting and critically revising the article for important intellectual content: Jorge Andrade-Sierra, Karla Hernández Morales, Carlos A Andrade Castellanos, Javier Arellano Arteaga, Andrade-Ortega Antonio de Jesús, Luis Gerardo González-Correa..
d. Final approval of the version to be published: All the team.

## Notes

### Competing Interest Statement

The authors have declared no competing interest.

### Funding Statement

This study did not receive any funding.

### Author Declarations

Ethics committee/IRB of Hospital Civil de Guadalajara, and Dr. Juan I Menchaca gave ethical approval for this work. Registration number: 17CI14 039 116 COFEPRIS

## REFERENCES

1. https://covid19.who.int/region/amro/country/mx.

2. Tian W, Jiang W, Yao J, Nicholson CJ, Li RH, Sigurslid HH, et al. Predictors of mortality in hospitalized COVID-19 patients: A systematic review and meta-analysis. J Med Virol. 2020;92(10):1875–83.

3. Bertsimas D, Lukin G, Mingardi L, Nohadani O, Orfanoudaki A, Stellato B, et al. COVID-19 mortality risk assessment: An international multi-center study. PLoS One. 2020;15(12):e0243262.

4. Cheng Y, Luo R, Wang K, Zhang M, Wang Z, Dong L, et al. Kidney disease is associated with in-hospital death of patients with COVID-19. Kidney Int. 2020;97(5):829–38.

5. Izzedine H, Jhaveri KD. Acute kidney injury in patients with COVID-19: an update on the pathophysiology. Nephrol Dial Transplant. 2021;36(2):224–6.

6. Pei G, Zhang Z, Peng J, Liu L, Zhang C, Yu C, et al. Renal Involvement and Early Prognosis in Patients with COVID-19 Pneumonia. J Am Soc Nephrol. 2020;31(6):1157–65.

7. Hirsch JS, Ng JH, Ross DW, Sharma P, Shah HH, Barnett RL, et al. Acute kidney injury in patients hospitalized with COVID-19. Kidney Int. 2020;98(1):209–18.

8. Moledina DG, Simonov M, Yamamoto Y, Alausa J, Arora T, Biswas A, et al. The Association of COVID-19 With Acute Kidney Injury Independent of Severity of Illness: A Multicenter Cohort Study. Am J Kidney Dis. 2021;77(4):490–9 e1.

9. Singh J, Malik P, Patel N, Pothuru S, Israni A, Chakinala RC, et al. Kidney disease and COVID-19 disease severity-systematic review and meta-analysis. Clin Exp Med. 2022;22(1):125–35.

10. Raina R, Mahajan ZA, Vasistha P, Chakraborty R, Mukunda K, Tibrewal A, et al. Incidence and Outcomes of Acute Kidney Injury in COVID-19: A Systematic Review. Blood Purif. 2022;51(3):199–212.

11. Ali H, Daoud A, Mohamed MM, Salim SA, Yessayan L, Baharani J, et al. Survival rate in acute kidney injury superimposed COVID-19 patients: a systematic review and meta-analysis. Ren Fail. 2020;42(1):393–7.

12. Lim JH, Park SH, Jeon Y, Cho JH, Jung HY, Choi JY, et al. Fatal Outcomes of COVID-19 in Patients with Severe Acute Kidney Injury. J Clin Med. 2020;9(6).

13. Diao B, Wang C, Wang R, Feng Z, Zhang J, Yang H, et al. Human kidney is a target for novel severe acute respiratory syndrome coronavirus 2 infection. Nat Commun. 2021;12(1):2506.

14. Abbate M, Rottoli D, Gianatti A. COVID-19 Attacks the Kidney: Ultrastructural Evidence for the Presence of Virus in the Glomerular Epithelium. Nephron. 2020;144(7):341–2.

15. Martinez-Rojas MA, Vega-Vega O, Bobadilla NA. Is the kidney a target of SARS-CoV-2? Am J Physiol Renal Physiol. 2020;318(6):F1454–F62.

16. Ronco C, Reis T. Kidney involvement in COVID-19 and rationale for extracorporeal therapies. Nat Rev Nephrol. 2020;16(6):308–10.

17. Nadim MK, Forni LG, Mehta RL, Connor MJ, Jr., Liu KD, Ostermann M, et al. COVID-19-associated acute kidney injury: consensus report of the 25th Acute Disease Quality Initiative (ADQI) Workgroup. Nat Rev Nephrol. 2020;16(12):747–64.

18. Joannidis M, Forni LG, Klein SJ, Honore PM, Kashani K, Ostermann M, et al. Lung-kidney interactions in critically ill patients: consensus report of the Acute Disease Quality Initiative (ADQI) 21 Workgroup. Intensive Care Med. 2020;46(4):654–72.

19. Ou M, Zhu J, Ji P, Li H, Zhong Z, Li B, et al. Risk factors of severe cases with COVID-19: a meta-analysis. Epidemiol Infect. 2020;148:e175.

20. Huang I, Pranata R, Lim MA, Oehadian A, Alisjahbana B. C-reactive protein, procalcitonin, D-dimer, and ferritin in severe coronavirus disease-2019: a meta-analysis. Ther Adv Respir Dis. 2020;14:1753466620937175.

21. Kermali M, Khalsa RK, Pillai K, Ismail Z, Harky A. The role of biomarkers in diagnosis of COVID-19 - A systematic review. Life Sci. 2020;254:117788.

22. Ciaccio M, Agnello L. Biochemical biomarkers alterations in Coronavirus Disease 2019 (COVID-19). Diagnosis (Berl). 2020;7(4):365–72.

23. Menez S, Moledina DG, Thiessen-Philbrook H, Wilson FP, Obeid W, Simonov M, et al. Prognostic Significance of Urinary Biomarkers in Patients Hospitalized With COVID-19. Am J Kidney Dis. 2022;79(2):257–67 e1.

24. Mahmudpour M, Roozbeh J, Keshavarz M, Farrokhi S, Nabipour I. COVID-19 cytokine storm: The anger of inflammation. Cytokine. 2020;133:155151.

25. Ragab D, Salah Eldin H, Taeimah M, Khattab R, Salem R. The COVID-19 Cytokine Storm; What We Know So Far. Front Immunol. 2020;11:1446.

26. Ronco C, Reis T, Husain-Syed F. Management of acute kidney injury in patients with COVID-19. Lancet Respir Med. 2020;8(7):738–42.

27. Levey AS, Stevens LA, Schmid CH, Zhang YL, Castro AF, 3rd, Feldman HI, et al. A new equation to estimate glomerular filtration rate. Ann Intern Med. 2009;150(9):604–12.

28. https://www.covid19treatmentguidelines.nih.gov/overview/clinical-spectrum/ [

29. Levey AS, Eckardt KU, Dorman NM, Christiansen SL, Hoorn EJ, Ingelfinger JR, et al. Nomenclature for kidney function and disease: report of a Kidney Disease: Improving Global Outcomes (KDIGO) Consensus Conference. Kidney Int. 2020;97(6):1117–29.

30. Lombardi R, Ferreiro A, Ponce D, Claure-Del Granado R, Aroca G, Venegas Y, et al. Latin American registry of renal involvement in COVID-19 disease. The relevance of assessing proteinuria throughout the clinical course. PLoS One. 2022;17(1):e0261764.

31. Casas-Aparicio GA, Leon-Rodriguez I, Alvarado-de la Barrera C, Gonzalez-Navarro M, Peralta-Prado AB, Luna-Villalobos Y, et al. Acute kidney injury in patients with severe COVID-19 in Mexico. PLoS One. 2021;16(2):e0246595.

32. Chavez-Iniguez JS, Cano-Cervantes JH, Maggiani-Aguilera P, Lavelle-Gongora N, Marcial-Meza J, Camacho-Murillo EP, et al. Mortality and evolution between community and hospital-acquired COVID-AKI. PLoS One. 2021;16(11):e0257619.

33. van Son J, Oussaada SM, Sekercan A, Beudel M, Dongelmans DA, van Assen S, et al. Overweight and Obesity Are Associated With Acute Kidney Injury and Acute Respiratory Distress Syndrome, but Not With Increased Mortality in Hospitalized COVID-19 Patients: A Retrospective Cohort Study. Front Endocrinol (Lausanne). 2021;12:747732.

34. Chan L, Chaudhary K, Saha A, Chauhan K, Vaid A, Zhao S, et al. AKI in Hospitalized Patients with COVID-19. J Am Soc Nephrol. 2021;32(1):151–60.

35. Fisher M, Neugarten J, Bellin E, Yunes M, Stahl L, Johns TS, et al. AKI in Hospitalized Patients with and without COVID-19: A Comparison Study. J Am Soc Nephrol. 2020;31(9):2145–57.

36. Moon H, Chin HJ, Na KY, Joo KW, Kim YS, Kim S, et al. Hyperphosphatemia and risks of acute kidney injury, end-stage renal disease, and mortality in hospitalized patients. BMC Nephrol. 2019;20(1):362.

37. Lin J, Chen J, Wu D, Li X, Guo X, Shi S, et al. Biomarkers for the early prediction of contrast-induced nephropathy after percutaneous coronary intervention in adults: A systematic review and meta-analysis. Angiology. 2022;73(3):207–17.

38. Feng Y, He H, Jia C, Xu Z, Li Y, Liao D. Meta-analysis of procalcitonin as a predictor for acute kidney injury. Medicine (Baltimore). 2021;100(10):e24999.

39. Chun K, Chung W, Kim AJ, Kim H, Ro H, Chang JH, et al. Association between acute kidney injury and serum procalcitonin levels and their diagnostic usefulness in critically ill patients. Sci Rep. 2019;9(1):4777.

40. Nie X, Wu B, He Y, Huang X, Dai Z, Miao Q, et al. Serum procalcitonin predicts development of acute kidney injury in patients with suspected infection. Clin Chem Lab Med. 2013;51(8):1655–61.

41. Dias BH, Rozario AP, Olakkengil SA, V A. Procalcitonin Strip Test as an Independent Predictor in Acute Pancreatitis. The Indian journal of surgery. 2015;77(Suppl 3):1012–7.

42. Simsek O, Kocael A, Kocael P, Orhan A, Cengiz M, Balci H, et al. Inflammatory mediators in the diagnosis and treatment of acute pancreatitis: pentraxin-3, procalcitonin and myeloperoxidase. Archives of Medical Science. 2018;14(2):288–96.

43. Level C, Chauveau P, Delmas Y, Lasseur C, Pelle G, Peuchant E, et al. Procalcitonin: a new marker of inflammation in haemodialysis patients? Nephrol Dial Transplant. 2001;16(5):980–6.

44. Nakamura Y, Murai A, Mizunuma M, Ohta D, Kawano Y, Matsumoto N, et al. Potential use of procalcitonin as biomarker for bacterial sepsis in patients with or without acute kidney injury. J Infect Chemother. 2015;21(4):257–63.

45. Amour J, Birenbaum A, Langeron O, Le Manach Y, Bertrand M, Coriat P, et al. Influence of renal dysfunction on the accuracy of procalcitonin for the diagnosis of postoperative infection after vascular surgery. Crit Care Med. 2008;36(4):1147–54.

46. Wang RR, He M, Kang Y. A risk score based on procalcitonin for predicting acute kidney injury in COVID-19 patients. J Clin Lab Anal. 2021;35(6):e23805.

47. Okusa MD. The inflammatory cascade in acute ischemic renal failure. Nephron. 2002;90(2):133–8.

48. Araujo M, Doi SQ, Palant CE, Nylen ES, Becker KL. Procalcitonin induced cytotoxicity and apoptosis in mesangial cells: implications for septic renal injury. Inflamm Res. 2013;62(10):887–94.

49. Spring JL, Winkler A, Levy JH. The influence of various patient characteristics on D-dimer concentration in critically ill patients and its role as a prognostic indicator in the intensive care unit setting. Clin Lab Med. 2014;34(3):675–86.

50. Zou X, Li S, Fang M, Hu M, Bian Y, Ling J, et al. Acute Physiology and Chronic Health Evaluation II Score as a Predictor of Hospital Mortality in Patients of Coronavirus Disease 2019. Crit Care Med. 2020;48(8):e657–e65.

51. Kashani K, Rosner MH, Haase M, Lewington AJP, O’Donoghue DJ, Wilson FP, et al. Quality Improvement Goals for Acute Kidney Injury. Clin J Am Soc Nephrol. 2019;14(6):941–53.

